# Psychological Variables Mediate Symptoms in Persistent Postural-Perceptual Dizziness (PPPD): A Cross-Sectional Self-Report Study

**DOI:** 10.1101/2024.10.19.24315702

**Authors:** Ariel Sereda, Ju Cheng Lam, Ali-Mert Hazar, Toby Ellmers, John Golding, Diego Kaski

## Abstract

**Background:** Persistent Postural-Perceptual Dizziness (PPPD) is a prevalent long-term functional neurological disorder characterised by non-spinning vertigo, perceived instability, and visual motion sensitivity. Current diagnostic criteria inadequately incorporate psychological variables widely associated with PPPD symptom onset and maintenance.

**Objectives:** This study explored PPPD-specific psychological variables to differentiate PPPD patients from healthy controls and, exploratorily, from Bilateral Vestibulopathy (BVP) patients. We evaluated these variables as potential treatment targets through mediation analysis. Our aim was to inform more precise diagnostic criteria and guide targeted interventions for PPPD.

**Methods:** We conducted a cross-sectional study with 164 participants, including 59 diagnosed cases of PPPD, 16 cases of BVP, and 89 healthy controls. Participants completed a series of questionnaires assessing negative illness perception, balance vigilance, anxiety, visual sensitivity, dizziness and other related metrics.

**Results:** Psychological variables, particularly anxiety, cognitive fusion, and justice appraisal significantly mediated the relationship between key PPPD symptoms (dizziness, visual sensitivity, and balance vigilance) and PPPD diagnosis compared to healthy controls. Logistic regression suggested psychological differences between PPPD and BVP, but limited BVP sample size constrained generalisability. Between PPPD and healthy controls, psychological variables significantly improved classification accuracy compared to measures of dizziness alone.

**Conclusion:** Incorporating psychological variables in the diagnosis and management of PPPD could enhance the understanding of the disorder and may aid in developing better-targeted interventions. The study supports revising existing diagnostic criteria to include validated psychological assessments and highlights the potential of treatments addressing cognitive and emotional aspects of PPPD to improve patient outcomes.

## Introduction

Recognised in 2017, Persistent Postural-Perceptual Dizziness (PPPD) is a chronic condition that severely disrupts daily activities with symptoms such as non-spinning vertigo, perceived instability, and psychological distress[1]. Patients often adopt maladaptive balance strategies such as stiffened gait and posture, exaggerated perceptions of movement including head tilt [2], postural sway [3] and low perceptual thresholds for vestibular motion [4]. Function is impaired by heightened self-monitoring, environmental vigilance, excessive compensatory behaviours in response to perceived or anticipated unsteadiness, hypervigilance towards vestibular and balance systems, reduced spatial cognition [5], and increased sway at the limit of stability [6]. PPPD often follows conditions like vestibular neuritis but can also occur without any vestibular injury [7]. Patients tend to prioritise visual stimuli over vestibular and somatosensory inputs, thought to alter vestibular responses [8]. Despite dependence on visual stimuli, patients exhibit high thresholds for detecting visual motion [9]. Clinical features of PPPD also vary with age. Younger patients (19-44 years) report higher anxiety, while older patients (65-85 years) often present with precipitating vestibular conditions [10].

Despite accounting for an estimated 20% of cases in neurology clinics, PPPD treatment lacks standardisation [11, 12]. As the leading cause of chronic neuro-otologic symptoms, investigating PPPD’s mechanisms to develop effective treatments is vital to improve patient outcomes [13]. Current diagnostic criteria (Table 1) do not include validated measures for psychological factors critical to PPPD, such as anxiety and negative illness perceptions [14, 15]. A Cognitive-Behavioural Model of PPPD based on Cognitive Behavioural Therapy (CBT) and Acceptance and Commitment Therapy (ACT) shows promise, but further work is needed to develop psychometric tools that capture relevant psychological variables[16–19].

**Table 1.** Barany Society Diagnostic Criteria for Persistent Postural-Perceptual Dizziness (PPPD)

| Criterion | Description |
| --- | --- |
| A | One or more symptoms of dizziness, unsteadiness, or non-spinning vertigo are present on most days for at least 3 months. |
| B | Symptoms of PPPD should occur without specific provocation, but are exacerbated by:<br>• Upright posture<br>• Active or passive motion<br>• Exposure to moving visual stimuli or complex visual patterns |
| C | Disorder is precipitated by conditions that cause vertigo, unsteadiness, dizziness, or problems with balance. Conditions may include acute, episodic, or chronic vestibular syndromes, other neurologic or medical illnesses, or psychological distress. |
| D | Symptoms of PPPD cause significant distress or functional impairment. |
| E | The symptoms of PPPD are not better explained by another disease or disorder. |
| *The Barany Society Diagnostic Criteria for PPPD includes five key aspects (criteria-A-E), and all five must be met to establish the diagnosis [1]* |  |

Current research relies heavily on non-specific patient-reported outcomes, often without biometric data, which limits the precision of treatment evaluation. For example, Herdman et al. (2022) demonstrated that redirecting attention from consciously controlling balance and fear-driven adaptations can significantly reduce negative illness perception. However, since negative illness perception is not part of PPPD’s diagnostic criteria, and the exact role of negative illness perception on dizziness is unknown, it is challenging to evaluate the effectiveness of this approach.

Developing psychometric tools to measure key psychological factors like negative cognitive appraisal and anxiety, could standardise assessments for PPPD and improve research and treatment.

## Objective

This study aims to bridge the gap between current diagnostic criteria and emerging understanding of PPPD by assessing psychological variables associated with its manifestation and maintenance. We evaluate these variables as potential treatment targets through mediation analysis between dizziness symptoms and PPPD diagnosis and investigate how they differentiate PPPD from healthy controls. We explore comparisons between PPPD, healthy controls (HC), and Bilateral Vestibulopathy patients (BVP) through multinomial logistic regression. We seek to inform more precise diagnostic criteria and guide the development of targeted psychological interventions.

## Method

### Design

A cross-sectional online questionnaire study. Participants provided electronic consent and completed 12 anonymised questionnaires via Qualtrics.

### Patients & Procedure

Adult patients diagnosed with PPPD and BVP from the University College London Hospital Vestibular Neurology Clinic were considered for this study. BVP was chosen as the disease control group due to its long-term symptoms of dizziness with a structural cause, contrasting with PPPD’s functional dizziness [20]. We recruited 195 participants between May to December 2023. Thirty-one were excluded due to co-occurring PPPD and BVP or ‘other vestibular disorder’. We included 164 participants (Mean age = 46.6 years, SD = 19.0, 106 females, and 58 males), 59 PPPD (mean age = 54.1 years, SD = 13.4), 16 BVP (mean age = 72.6 years, SD = 10.0), and 89 HC (mean age = 36.9 years, SD = 16.7). The average disease duration was 4.0 years, for PPPD (mean 3.9 years, SD = 1.6) and for BVP (mean 4.5 years, SD = 1.4). Ethics approval was obtained from the UCL/UCLH Joint Research Office and the University of Leicester Research Ethics Committee.

### Measures

To assess vulnerability factors and clinically significant symptoms outlined in both the Barany Society Diagnostic Criteria [1] and the Cognitive-Behavioural Model of PPPD [16] we included psychological questionnaires to measure maladaptive changes in cognitive appraisal, emotional response, attention, and behaviour (Figure 1). Where applicable, subscales and clinical cutoffs were accounted for (Table 2).

**Figure 1.**
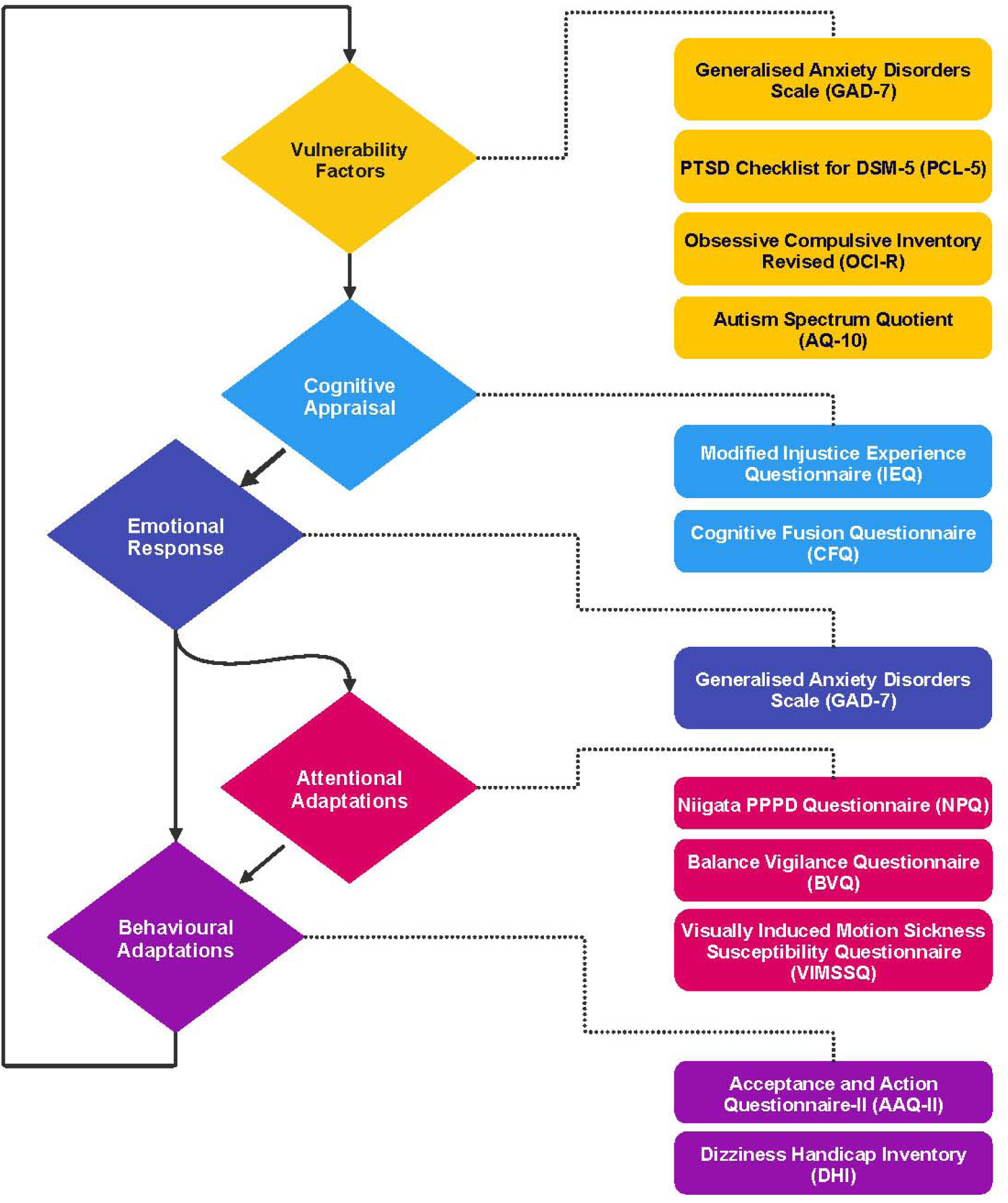
Combined Model of PPPD with Associated Psychometric Questionnaires. Generalised Anxiety Disorders Scale (GAD-7), PTSD Checklist for DSM-5 (PCL-5), Modified Injustice Experience Questionnaire (IEQ), Cognitive Fusion Questionnaire (CFQ), Balance Vigilance Questionnaire (BVQ), Visually Induced Motion Sickness Susceptibility Questionnaire (VIMSSQ), The Acceptance and Action Questionnaire-II (AAQ-II), Dizziness Handicap Inventory (DHI), Niigata PPPD Questionnaire (NPQ).

**Table 2.** Questionnaires Included.

| **Questionnaire** | **Items** | **Individual Item Scoring range** | **Cutoff** | **Highest Score** | **Subscales** |
| --- | --- | --- | --- | --- | --- |
| Niigata PPPD Questionnaire (NPQ) | 12 | 0-6 | 27 | 72 | • Visual<br>• Active<br>• Passive |
| Dizziness Handicap Inventory (DHI) | 25 | • Yes<br>• Sometimes<br>• No<br>• N/A | 10 | 100 | • Physical<br>• Functional<br>• Emotional |
| Visually Induced Motion Sickness Susceptibility Questionnaire (VIMSSQ) | 6 | • Never<br>• Rarely<br>• Sometimes<br>• Often | None | 18 | None |
| Balance Vigilance Questionnaire (BVQ) | 6 | 1-5 | 19 | 30 | None |
| Generalised Anxiety Disorder Scale (GAD7) | 7 | 0-3 | 8 | 21 | None |
| Modified Injustice Experience Questionnaire, IEQ-chr, (IEQ) | 12 | 0-4 | 19 | 48 | • Severity/Irreparability<br>• Blame/Unfairness |
| Cognitive Fusion Questionnaire (CFQ) | 7 | 1-7 | None | 49 | None |
| The Acceptance and Action Questionnaire-II (AAQII) | 7 | 1-7 | 28 | 49 | None |
| PTSD Checklist for DSM-5 (PCL-5) | 20 | 0-4 | 31 | 80 | None |
| Obsessive-Compulsive Inventory (OCI-R) | 18 | 0-4 | 12 | 72 | None |
| Autism-Spectrum Quotient-10 (AQ10) | 10 | • Definitely Agree<br>• Slightly Agree<br>• Slightly Disagree<br>• Definitely Disagree | 6 | 10 | None |

The Generalised Anxiety Disorder Scale (GAD-7) assesses anxiety levels [21], a key vulnerability factor and emotional response influencing PPPD onset and progression [14]. The PTSD Checklist for DSM-5 (PCL-5) measures post-traumatic stress-related symptoms that could worsen or trigger PPPD [22].

Improved illness perception is associated with recovery in PPPD patients [17]. The Modified Injustice Experience Questionnaire (IEQ) measures negative cognitive appraisal towards illness, linked to worse outcomes and slower recovery in chronic pain patients [23]. The Cognitive Fusion Questionnaire (CFQ) measures the degree of fusion with cognitive processes, a critical factor guiding the use of ACT techniques in treatment [24]. The Acceptance and Action Questionnaire-II (AAQ-II) assesses experiential avoidance [25]. Together, the CFQ and AAQ-II measure psychological flexibility, a primary measure in ACT and CBT, predictive of treatment effectiveness [26].

The Balance Vigilance Questionnaire (BVQ) measure attentional strategies towards balance, offering insights into how patients cope with balance challenges [27].

The Niigata PPPD Questionnaire (NPQ) assess behavioural adaptations (i.e. avoidance) and the impact of provocative situations on dizziness symptoms [28]. The Dizziness Handicap Inventory (DHI) measures the self-perceived impact of dizziness on emotional, physical, and functional subscales [29].

### Statistical Analysis

Data processing and analysis were done in Python 3.9 using Pandas, NumPy, SciPy, scikit-learn, Seaborn, and Statsmodels. Questionnaire responses were converted to numerical scores and normalized/standardized with MinMaxScaler and StandardScaler. Outliers were removed using the Interquartile Range (IQR) method.

Normality was assessed with the Shapiro-Wilk test. Group differences (HC, PPPD, BVP) were analysed using the Kruskal-Wallis test, with Dunn’s post-hoc test for pairwise comparisons. Effect sizes (epsilon-squared, Cliff’s Delta) and Holm-Bonferroni adjustments were applied. Pearson or Spearman methods calculated correlation coefficients.

Chi-square tests (or Fisher’s exact test) evaluated differences in clinically significant scores, with effect sizes using Cramer’s V or odds ratios.

Multinomial logistic regression classified diagnoses, assessing multicollinearity via the Variance Inflation Factor (VIF) and handling class imbalance with Synthetic Minority Over-sampling Technique (SMOTE). The model was evaluated using 5-fold cross-validation and performance metrics like precision, recall, and F1-score. Mediation analysis was done via logistic regression and OLS regression, with bootstrapping (1000 iterations) for confidence intervals.

### Supplementary Analysis

Supplementary materials provide additional analyses to support our findings. These include descriptive statistics (Figure S1) and ranked correlation coefficients (Figures S2-S3). Chi-square results comparing clinically significant results across groups (Table S1). Kruskal-Wallis results, Dunn’s post-hoc test for pairwise comparisons between groups with Epsilon-squared and Cliff’s Delta effect size measures (Table S2). Materials for mediation analysis (Figure S4, Table S3) and multinomial logistic regression (Figure S5, Table S2) include variance inflation factors and detailed results tables for both analyses.

## Results

### Key Psychological Findings in PPPD

Comparing the proportional group clinical cutoff scores underscored the burden of psychological and dizziness factors in PPPD and BVP patients compared to healthy controls (Figure 2). Chi-square tests (or Fisher’s exact test when frequency was below 5) showed significant differences amongst groups for most questionnaires, excluding OCI and AQ10. After adjusting for multiple comparisons, PPPD patients showed notably higher anxiety (GAD-7) and experiential avoidance (AAQ-II) scores, whilst BVP patients exhibited a unique pattern of balance-related vigilance (BVQ). Strong effect sizes were observed between PPPD and HC groups across several measures. However, the small BVP sample size warrants cautious interpretation. The findings suggest that anxiety and experiential avoidance may be key differentiating factors between PPPD and BVP, whereas obsessive-compulsive traits and autistic-like characteristics appear less distinctive across these patient populations.

**Figure 2.**
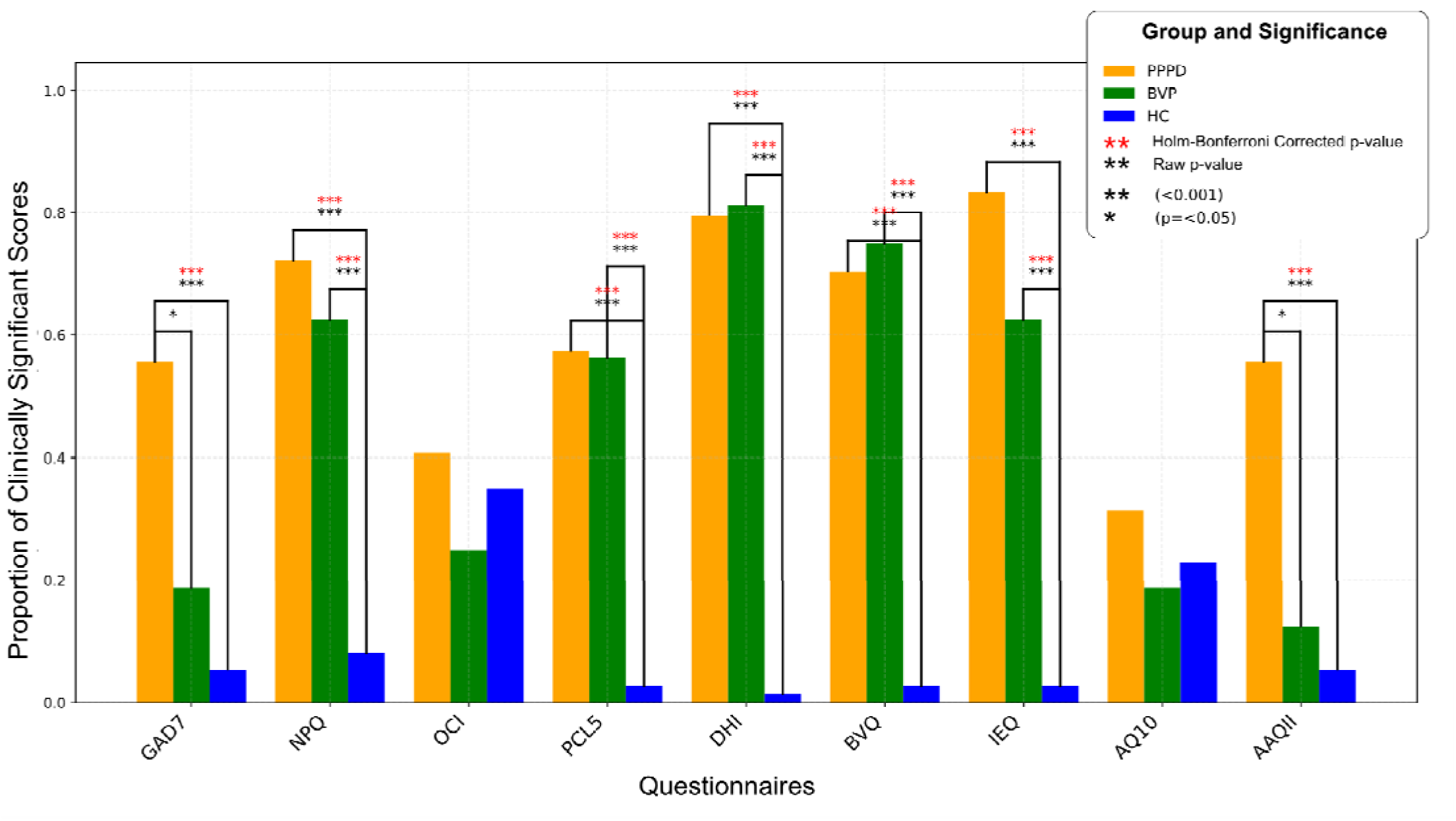
Proportion of Clinically Significant Scores by Group and Questionnaire. Bar graph displays the proportion of clinically significant scores across different questionnaires for three groups: PPPD (Persistent Postural-Perceptual Dizziness), BVP (Bilateral Vestibulopathy), and HC (Healthy Controls). Each bar represents the proportion of participants in each group who scored above a clinically significant cutoff on a specific questionnaire. The cutoffs for clinical significance are indicated below each questionnaire on the x-axis. PPPD displayed proportionally higher scores compared to HC and BVP groups for all but one questionnaire (BVQ), where BVP recorded the highest proportion of significant scores. Statistically significant differences between groups (p < 0.05) were observed for GAD-7, NPQ, PCL-5, DHI, BVQ, IEQ, and AAQ-II, as determined by 3×2 Chi-Square tests. No significant differences were found for OCI and AQ10. Pairwise comparisons revealed varying effect sizes (Cramer’s V) and odds ratios for significant differences between specific group pairs. *Questionnaires with clinical significance cutoffs: Generalised Anxiety Disorders Scale (GAD-7), Niigata PPPD Questionnaire (NPQ), Obsessive Compulsive Inventory (OCI), PTSD Checklist for DSM-5 (PCL-5), Dizziness Handicap Inventory (DHI), Balance Vigilance Questionnaire (BVQ), Modified Injustice Experience Questionnaire (IEQ), Autism-Spectrum-Quotient (AQ10), The Acceptance and Action Questionnaire-II (AAQ-II).

Proportional cutoff scores between PPPD and BVP were most similar on the dizziness handicap inventory (DHI). Although BVQ scores in BVP compared to PPPD were generally higher (as indicated by the odds ratio being less than 1), the difference is insignificant after correcting for multiple comparisons.

### Comparative Analysis of Questionnaire Scores

Kruskal-Wallis ANOVA tests reveal that PPPD and BVP differ significantly from HC across almost all measures, indicating that these conditions are distinct from the general healthy population (Figure 3).

**Figure 3.**
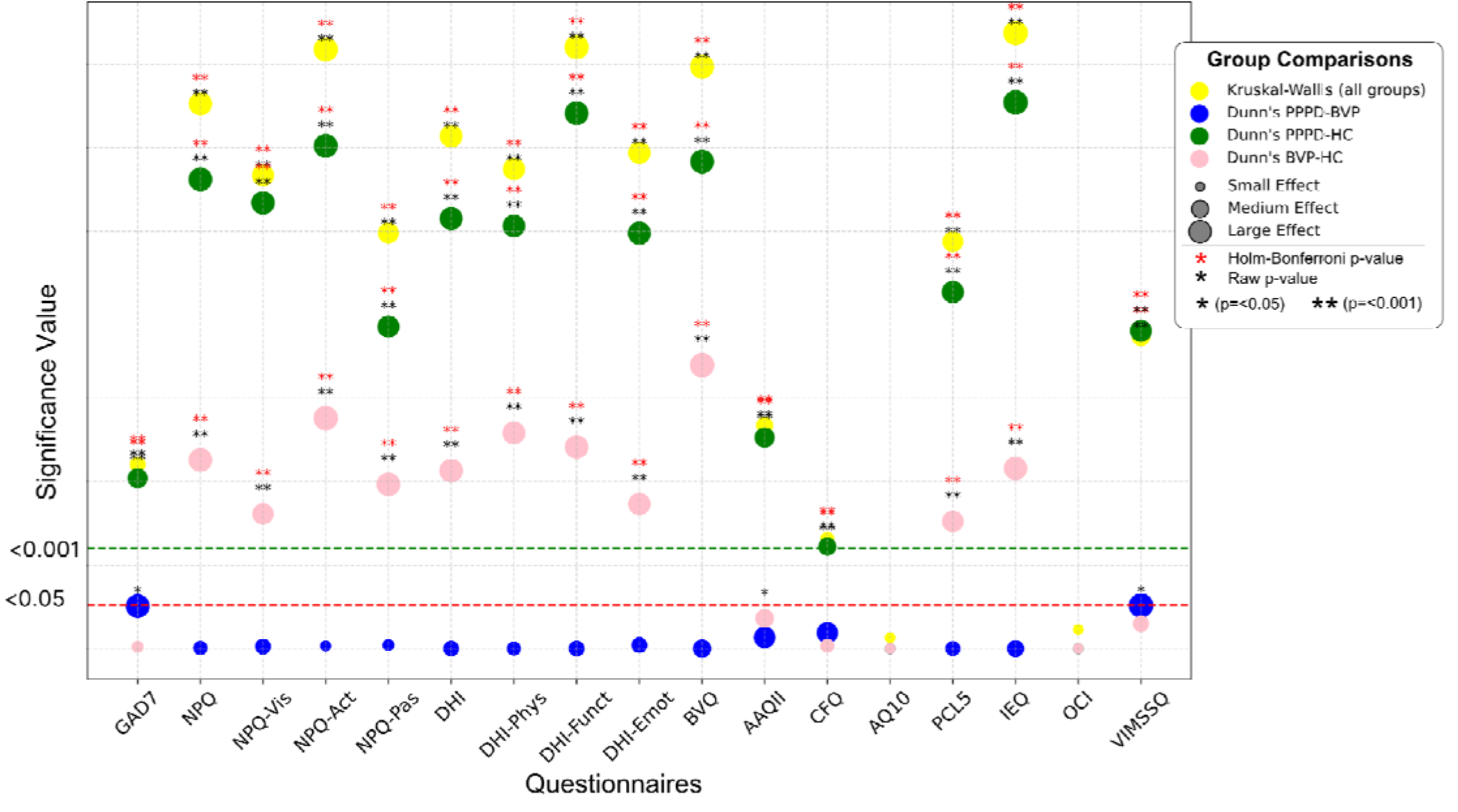
Significance Levels for Differences in Selected Questionnaire Scores Across Groups. Dot plot displaying p-values and effect sizes for group comparisons (Kruskal-Wallis test for overall group differences, and Dunn’s test for pairwise comparisons: PPPD vs BVP, PPPD vs HC, BVP vs HC) across various questionnaires (for ease of visualisation, p-values are transformed using -log10). The horizontal dashed lines indicate the significance thresholds for p = 0.05 (red) and p = 0.001 (green). Marker size represents effect size, with larger markers indicating stronger effects. Significance is denoted by asterisks: * (p ≤ 0.05) and **(p ≤ 0.001), with black asterisks for raw p-values and red asterisks for Holm-Bonferroni corrected p-values. The questionnaires are as follows: Generalised Anxiety Disorders Scale (GAD-7), Niigata PPPD Questionnaire (NPQ), NPQ-Visual (NPQ-Vis), NPQ-Active (NPQ-Act), NPQ-Passive (NPQ-Pas), Dizziness Handicap Inventory (DHI), DHI-Physical, DHI-Functional, DHI-Emotional, Balance Vigilance Questionnaire (BVQ), The Acceptance and Action Questionnaire-II (AAQ-II), Cognitive Fusion Questionnaire (CFQ), Autism-Spectrum Quotient-10 (AQ10), PTSD Checklist for DSM-5 (PCL-5), Modified Injustice Experience Questionnaire (IEQ), Obsessive-Compulsive Inventory (OCI), and Visually Induced Motion Sickness Susceptibility Questionnaire (VIMSSQ).

Differences across most questionnaires were observed between groups, particularly between PPPD/BVP and healthy control groups. The largest effect sizes for overall group differences (Kruskal-Wallis epsilon-squared) were noted in the IEQ (ε^2^ = 0.588), DHI-Functional (ε^2^ = 0.574), NPQ-Active (ε^2^ = 0.572), and BVQ (ε^2^ = 0.555). The PPPD and BVP groups showed similarity in many measures, with fewer significant differences after Holm-Bonferroni correction. However, potential distinguishing features between PPPD and BVP include anxiety (GAD-7) and visually induced motion sickness (VIMSSQ), with notable effect sizes: GAD-7 (Cliff’s δ = 0.394), VIMSSQ (Cliff’s δ = 0.419), although these differences did not reach statistical significance after correction (p = 0.053 and p = 0.051, respectively).

AQ10 and OCI showed no significant differences between any groups.

### Psychological Variables Mediate Dizziness and PPPD Diagnosis

The mediation analysis (Figures 4 and 5) comparing PPPD diagnoses to healthy controls revealed that anxiety (GAD-7), perceived injustice (IEQ), and cognitive fusion (CFQ) all significantly mediate the relationship between attentional and behavioural dizziness symptom questionnaires (NPQ, BVQ, and VIMSSQ) and PPPD diagnosis.

**Figure 4.**
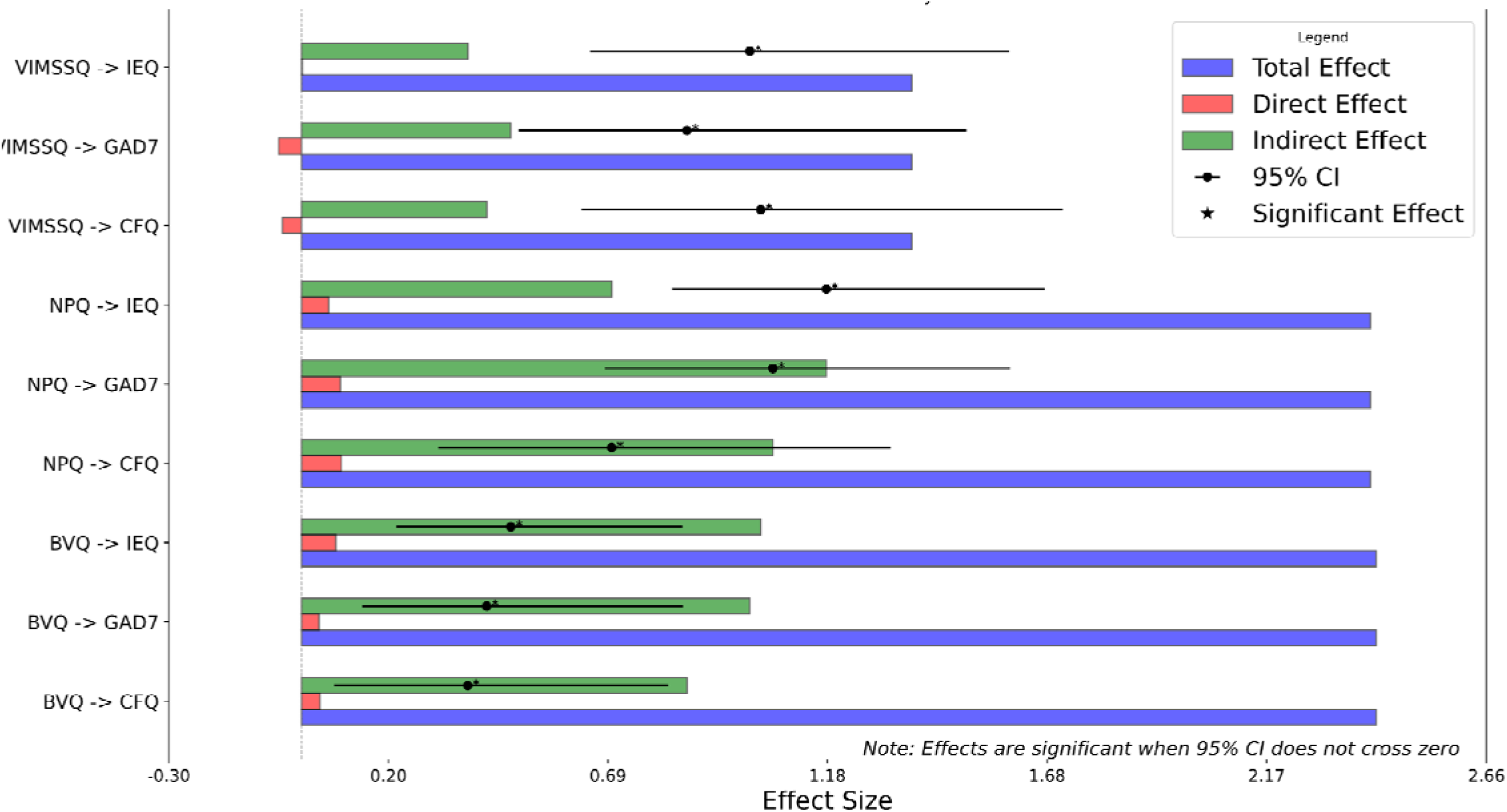
Mediation Analysis Results by Significance and Strength of Indirect Effect of Justice Appraisal (IEQ) and Anxiety (GAD-7) on Dizzy Symptoms and PPPD Diagnosis. This figure shows total, direct, and indirect effects for each pathway, with 95% confidence intervals. Effects are considered significant when the confidence interval does not cross zero. Notably, the model reveals small and negative direct effects of NPQ, BVQ, and VIMSSQ on PPPD diagnosis. A negative direct effect indicates that when controlling for mediators (e.g., anxiety, perceived injustice), the independent variable (IV), such as NPQ or VIMSSQ, has a suppressive or negative influence on the dependent variable (DV, in this case, PPPD diagnosis). This suggests that the relationship between these dizziness measures and PPPD is primarily explained through the mediating psychological variables rather than a direct influence.

**Figure 5:**
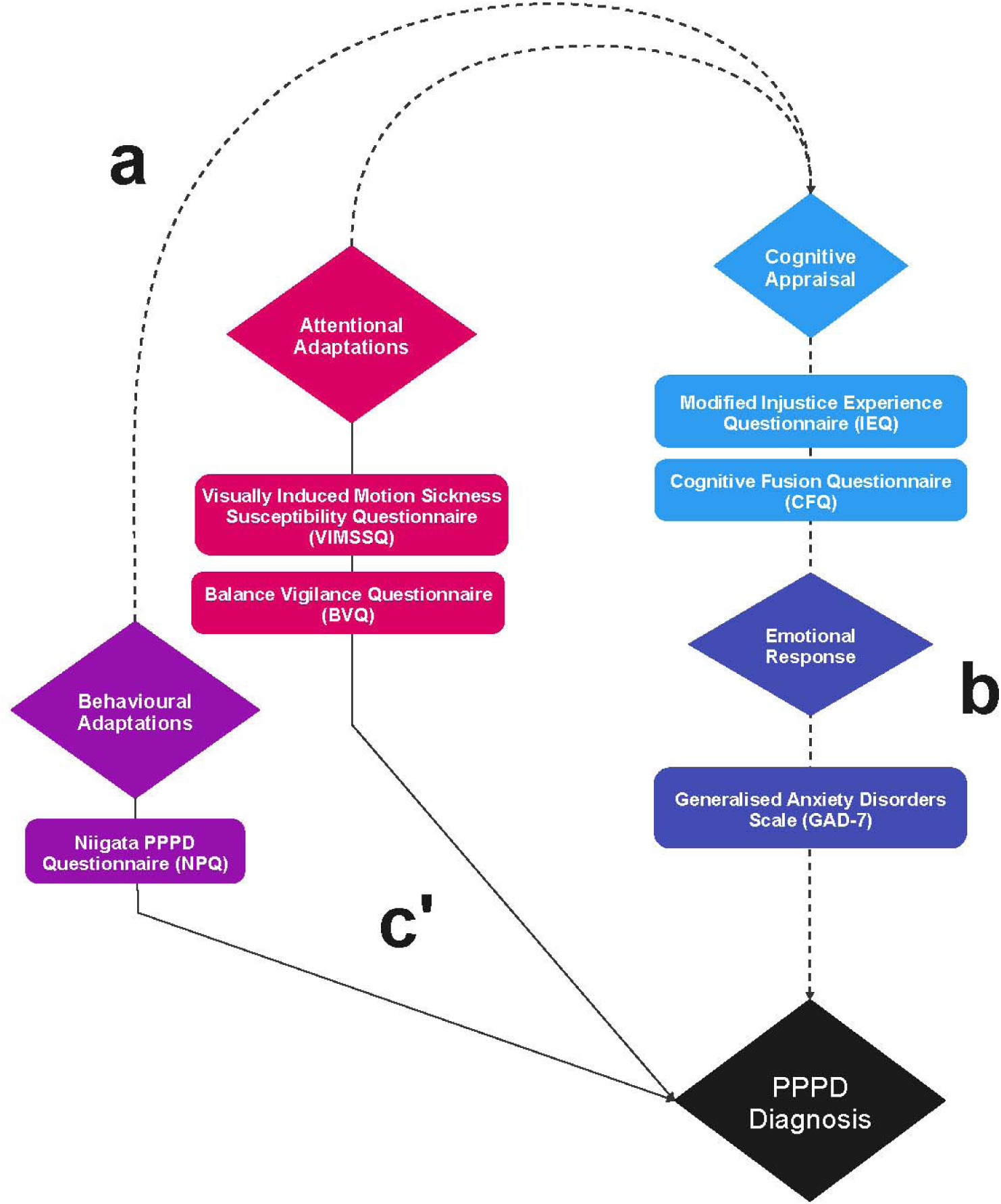
Path Diagram Illustrating Mediation Effects on PPPD Diagnosis. This diagram shows the mediation analysis of the pathways leading to the diagnosis of Persistent Postural-Perceptual Dizziness (PPPD). The independent variables (IVs) include the Niigata PPPD Questionnaire (NPQ), Visually Induced Motion Sickness Susceptibility Questionnaire (VIMSSQ), and the Balance Vigilance Questionnaire (BVQ). These IVs are mediated through the Modified Injustice Experience Questionnaire (IEQ), Cognitive Fusion Questionnaire (CFQ), and the Generalised Anxiety Disorders Scale (GAD-7), representing cognitive appraisal and emotional response, respectively. The pathways (a) illustrate the effect of IVs on mediators, (b) show the effect of mediators on the dependent variable (DV), and (c’) represent the direct effect of IVs on the diagnosis of PPPD. The mediation analysis results demonstrate significant indirect effects, with small and negative direct effects, highlighting the critical roles of anxiety, perceived injustice, and cognitive fusion in the diagnosis of PPPD. These negative direct effects suggest that the influence of dizziness-related questionnaires on PPPD diagnosis is primarily explained through the mediators, rather than a direct relationship.

Notably, the VIMSSQ exhibited small and negative direct effects on PPPD diagnosis when accounting for the mediators, indicating suppressor effects, suggesting that the influence of visually induced motion sickness susceptibility on PPPD diagnosis operates primarily through the psychological mediators of anxiety, cognitive fusion, and perceived injustice, rather than through a direct relationship.

In contrast, the NPQ and BVQ showed small but positive direct effects, indicating that these questionnaires capture symptoms directly associated with PPPD diagnosis, independent of psychological mediators

The NPQ significantly impacts the diagnosis of PPPD through all three mediators, anxiety (GAD-7), perceived injustice (IEQ), and cognitive fusion (CFQ). The total effect of NPQ on PPPD diagnosis is substantial at 2.421, with the direct effect being minimal at 0.060. The indirect effects through anxiety (GAD-7) (1.157, CI: [0.809, 1.687]), perceived injustice (IEQ) (0.748, CI: [0.327, 1.377]), and cognitive fusion (CFQ) (1.054, CI: [0.679, 1.613]) are all significant, highlighting the critical roles these psychological factors play in mediating the relationship between NPQ scores and PPPD diagnosis.

Similarly, the BVQ exhibits a significant impact on PPPD diagnosis through anxiety (GAD-7), perceived injustice (IEQ), and cognitive fusion (CFQ). The total effect is the highest among the questionnaires at 2.417, with significant indirect effects through GAD-7 (1.056, CI: [0.620, 1.775]), IEQ (1.056, CI: [0.620, 1.775]), and CFQ (0.875, CI: [0.500, 1.462]). The direct effect remains small and negative, suggesting that these mediators are essential in explaining the relationship between BVQ scores and PPPD diagnosis.

Both NPQ and BVQ have small positive direct effects on PPPD diagnosis when accounting for the mediators, indicating that while the mediators significantly contribute to the relationship, there is still a small direct effect of these questionnaires on PPPD diagnosis.

In contrast, the VIMSSQ shows small and negative direct effects on PPPD diagnosis when accounting for the mediators, anxiety (GAD-7) and cognitive fusion (CFQ), while perceived injustice (IEQ) showed no direct effect. The small negative direct effects observed with VIMSSQ when mediated by GAD-7 and CFQ suggest suppressor effects. A suppressor effect occurs when the inclusion of a mediator increases the predictive validity of an independent variable by accounting for variance that suppresses the direct relationship. This implies that the direct relationship between VIMSSQ and PPPD diagnosis is weak or negative, and the association is primarily explained through the mediating psychological variables of anxiety and cognitive fusion.

The total effect of VIMSSQ on PPPD diagnosis is 1.372, indicating a strong overall association between VIMSSQ scores and PPPD diagnosis. Finally, the indirect effects through GAD-7 (0.470, CI: [0.212, 0.852]) and CFQ (0.416, CI: [0.133, 0.892]) are significant, highlighting the roles of anxiety and cognitive fusion. The indirect effect through IEQ is smaller (0.374, CI: [0.050, 0.840]) and marginally significant, suggesting that perceived injustice does not play a substantial mediating role between VIMSSQ scores and PPPD diagnosis. The presence of suppressor effects with the VIMSSQ underscores the importance of these psychological mediators in explaining the influence of visually induced motion sickness susceptibility on PPPD diagnosis.

### Questionnaires for Differential Diagnosis

Multinomial logistic regression predicted group membership between PPPD, BVP, and HC. The model included six questionnaires: dizziness (NPQ), visually induced motion sickness (VIMSSQ), balance vigilance (BVQ), anxiety (GAD-7), PTSD (PCL-5) and injustice experience (IEQ). Age and gender were included as controls. All predictors were standardised for comparability. A VIF cutoff of 5 addressed multicollinearity among predictors. The Dizziness Inventory (DHI) was excluded due to high multicollinearity with the NPQ (VIF of 7).

Overall PPPD classification had a precision, recall, and F1-score of 0.875; HC had a precision of 0.917, recall of 0.957, and F1-score of 0.936; BVP, had a precision of 0.750, recall of 0.600, and F1-score of 0.667. The overall model accuracy was 88.64%, with a pseudo R-squared value of 0.7414 (variance explained by the model). Given the small BVP sample size, results are preliminary and warrant further investigation with larger samples.

Pairwise classification between PPPD and HC reported 92% accuracy, with a precision of 0.933 and recall of 0.875 for PPPD, and a precision of 0.957 and recall of 0.957 for HC. For PPPD vs. BVP, the model achieved an accuracy of 81%, with a precision of 0.933 and recall of 0.875 for PPPD, and a precision of 0.750 and recall of 0.600 for BVP.

Standardised coefficients for Diagnosis=1 (PPPD vs. HC) showed significant predictors like IEQ (β = −2.473, p = 0.009). For Diagnosis=2 (PPPD vs. BVP), significant predictors included BVQ (β = 1.934, p = 0.031), VIMSSQ (β = −1.450, p = 0.010), PCL-5 (β = 1.701, p = 0.019), and Age (β = 14.381, p < 0.001). Positive beta coefficients suggest increases in the predictor variable indicate a higher outcome likelihood. For example, higher BVQ scores (β = 1.934) indicate increased likelihood of BVP over PPPD. Conversely, negative beta coefficients suggest increases in the predictor variable indicate lower likelihood, i.e. higher IEQ score (β = −2.473) decreases the likelihood of HC over PPPD. Standardised beta coefficients show changes in log-odds for a one standard deviation change in the predictor variables.

Odds ratios were calculated to complement the standardised coefficients to interpret the predictor’s effect on group membership probabilities. For PPPD vs HC, a one standard deviation increases in IEQ score reduced the odds of being in the HC group by 91.6% (OR = 0.084, p = 0.009). In the PPPD vs BVP comparison, significant predictors included BVQ (OR = 6.919, p = 0.031), VIMSSQ (OR = 0.235, p = 0.010), PCL-5 (OR = 5.477, p = 0.019), and age (OR = 1,762,042.837, p < 0.001).

**Figure 6.**
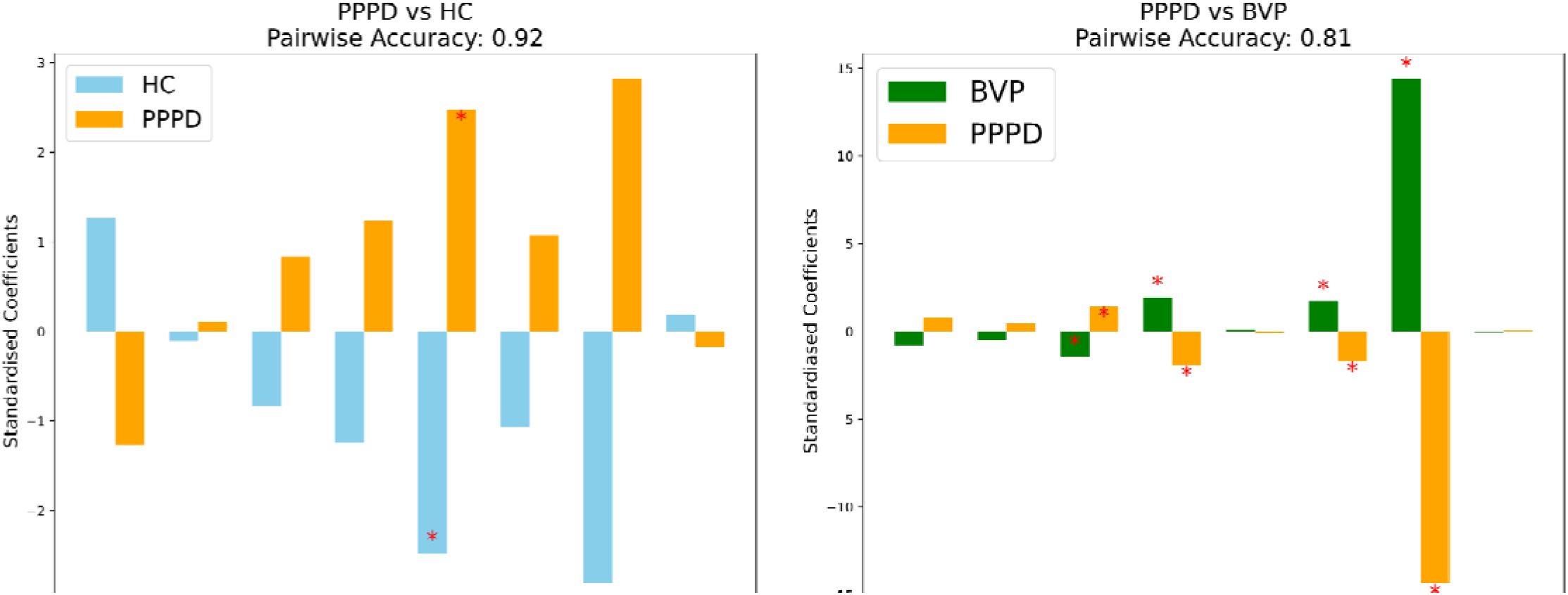
Multinomial Logistic Regression Coefficient Significance for PPPD vs. HC and PPPD vs. BVP. The left plot displays the standardised regression coefficients comparing Persistent Postural-Perceptual Dizziness (PPPD) to Healthy Controls (HC). Significant predictors are indicated by the red asterisk. Higher IEQ scores are associated with a higher likelihood of being in the PPPD group compared to the HC group. The right plot displays the standardised regression coefficients comparing PPPD to BVP (Bilateral Vestibulopathy). Higher BVQ and PCL-5 scores are associated with a higher likelihood of BVP compared to PPPD. Higher VIMSSQ scores are associated with a higher likelihood of being in the PPPD group compared to the BVP group.

Cross-validation using a 5-fold method on the dataset revealed a mean accuracy of 0.83. Accuracies across folds were 0.862, 0.897, 0.793, 0.724, and 0.893. Cross-validation log losses of 0.459, 0.371, 0.535, 0.760, and 0.286 evaluated how predicted probabilities matched the actual class labels. The mean log loss of 0.480 suggests fairly accurate probabilistic predictions and reasonable model performance.

The overall model accuracy of 88.64% demonstrates effectiveness in correctly predicting unseen data. The confusion matrix showed that out of 23 HC cases, the model correctly classified 22, with one misclassification. For PPPD, the model correctly identified 14 of 16 cases, with two misclassified as HC. For BVP, the model correctly classified 3 of 5 cases, with one misclassified as PPPD and one as HC. While these results are promising, they should be considered as leads for future research with larger BVP samples.

## Discussion

This study aimed to identify psychological variables important to PPPD by comparing patients with healthy controls and those with BVP. Through mediation analyses and multinomial logistic regression, we assessed interactions between psychological factors and dizziness symptoms.

Our mediation analysis shows that psychological factors, particularly perceptions, interpretations, and emotional responses, strongly influence PPPD symptom severity. Negative illness appraisal (IEQ), cognitive fusion (CFQ), and anxiety (GAD-7) mediated the relationship between dizziness symptoms (measured by NPQ, BVQ, and VIMSSQ) and PPPD diagnosis. This suggests that symptoms like dizziness, body vigilance, and visually induced motion sickness are strongly shaped by negative cognitive appraisals and heightened anxiety.

Strong total effects of the NPQ and BVQ on PPPD diagnosis, along with small direct effects, suggest these tools capture PPPD symptoms independent of psychological mediators. Yet, significant indirect effects via anxiety, cognitive fusion, and perceived injustice underscore the influence of these psychological factors.

Conversely, the VIMSSQ showed small negative direct effects on PPPD diagnosis after adjusting for mediators, indicating suppressor effects. Despite this, significant indirect effects through psychological mediators were observed. In patients with higher susceptibility to visually induced motion sickness, the impact on PPPD diagnosis appears to be primarily mediated by psychological factors rather than direct effects. This is notable given the emphasis on visually induced dizziness in PPPD [30].

Our findings indicate that visual motion sensitivity in PPPD is significantly influenced by psychological factors, not solely a sensory phenomenon. The suppressor effects observed with the VIMSSQ imply that psychological factors mediate the relationship between visual motion sensitivity and PPPD diagnosis, underscoring the necessity of addressing psychological factors in PPPD management.

Results from our multinomial logistic regression align with the accentuated role of visual dependence and sensitivity in PPPD [8, 14, 31], where the VIMSSQ was a key predictor distinguishing PPPD from BVP. Conversely, higher BVQ scores significantly predicted BVP diagnosis, suggesting that visual motion sensitivity, heavily influenced by psychological factors, may distinguish PPPD from structural causes of dizziness like BVP.

The overlap of NPQ and BVQ scores in PPPD and BVP underscores the importance of a personalised assessment approach, addressing both physical symptoms and associated psychological factors. The NPQ’s inability to significantly differentiate group membership suggests limitations in its diagnostic specificity; 75% of BVP patients scored above the PPPD clinical cutoff, highlighting the need for improved assessment tools that isolate functional and psychological factors important for PPPD.

High multicollinearity between the DHI and NPQ (VIF of 7) raises concerns about the distinctiveness of these instruments, potentially compromising their utility and the validity of analyses. This is problematic since the DHI includes an emotional subscale while the NPQ does not measure emotional aspects. Our results align with an independent validation of the NPQ, showing lower diagnostic accuracy (sensitivity 72%, specificity 56%) than initially reported (82%, 74%) [32]. The NPQ’s focus on exacerbating factors without psychological aspects is notable since the Bárány Society criteria specify psychological distress and significant distress in criteria C and D [1]. Our research supports expanding the current diagnostic criteria to include more precise psychological variables, such as PPPD-specific evaluations of cognitive appraisal and autonomic arousal.

Though findings are limited by sample size, almost all BVP patients scored above the cutoff on the BVQ [27], with average scores exceeding those of PPPD patients, suggesting that balance hypervigilance may be a generalised outcome of vestibular dysfunction. A higher proportion of PPPD patients exhibited clinically significant anxiety (49.02%) compared to BVP patients (28.57%), confirming prior observations [33]. Further study limitations include the cross-sectional design, which precludes causal inferences, and reliance on self-reported measures, which may introduce bias. Unmeasured variables like depression could have influenced results; controlling for these in future research would enhance robustness. The small BVP sample limits generalisability. Larger samples are needed to investigate the role of visually induced dizziness in differential diagnosis between PPPD and BVP.

Our results align with research underscoring the role of visual dependence, negative cognitive appraisal, and sympathetic arousal in PPPD. Studies show reduced functional connectivity in areas associated with multisensory vestibular processing and spatial cognition [34], and increased inter-network connectivity suggests visual information plays a dominant compensatory role. Visual arousal at rest shows altered FC patterns indicating compensatory reliance on visual and somatosensory inputs, and post-stimulus changes reveal enhanced connectivity within vestibular and visual networks but disruptions in somatosensory-visual circuits [35], reflecting challenges in sensory integration and emotional regulation [36].

Negative cognitive appraisal is a critical factor in PPPD, involving negative beliefs about illness [17, 18], and interpretations of sensory perceptions [2, 3, 8, 9]. Developing psychometric items to capture the range of negative cognitive appraisals in PPPD, focusing on general negative illness perceptions and the cognitive appraisal of specific perceptual scaling of sensory stimuli, is recommended.

Although the link between PPPD patient-reported anxiety and visual-vestibular interaction is established [37], the role of sympathetic autonomic arousal during physical movement remains underexplored [38]. In healthy individuals, vestibular self-motion perception becomes more intense under threat [39], possibly due to shared neural substrates with emotional processing. While catastrophic thinking and low resilience predict PPPD [14, 15], the mechanisms by which negative thoughts affect autonomic arousal, vestibular, and visual perceptual scaling in PPPD remain unknown [13].

Future research should develop PPPD-specific patient-reported and biometric tools to better understand the psychological and physical interactions in the condition. These outcome measures will support comprehensive treatment evaluation and clarify interactions between autonomic arousal, aberrant perceptual scaling and sensory thresholds [4, 5, 40] and attentional and behavioural adaptations in PPPD.

Our findings suggest incorporating psychological variables in PPPD diagnosis and management could enhance understanding and enable targeted interventions addressing both physical symptoms and psychological factors. For example, CBT targeting maladaptive thought patterns and emotional responses could manage anxiety and reduce cognitive fusion, potentially mediating visual motion sensitivity.

In conclusion, our findings support a combined cognitive-behavioural model of PPPD, demonstrating how cognitive appraisal and emotional response contribute to maladaptive attentional and behavioural strategies related to dizziness. Developing PPPD-specific outcome measures, including patient-reported and biometric assessments, will enable treatment evaluations. Mediation analysis suggests that interventions targeting cognitive reappraisal and autonomic arousal regulation could reduce the impact of negative cognitive appraisals, anxiety, and avoidance behaviours due to dizziness in PPPD. Future research should investigate the interplay between autonomic arousal, cognitive appraisal, and symptom severity.

## Data Availability

All data produced in the present study are available upon reasonable request to the authors.

